# Racial disparities in COVID-19 mortality are driven by unequal infection risks

**DOI:** 10.1101/2020.09.10.20192369

**Authors:** Jon Zelner, Rob Trangucci, Ramya Naraharisetti, Alex Cao, Ryan Malosh, Kelly Broen, Nina Masters, Paul Delamater

## Abstract

**Background:** As of August 5, 2020, there were more than 4.8M confirmed and probable cases and 159K deaths attributable to SARS-CoV-2 in the United States, with these numbers undoubtedly reflecting a significant underestimate of the true toll. Geographic, racial-ethnic, age and socioeconomic disparities in exposure and mortality are key features of the first and second wave of the U.S. COVID-19 epidemic.

**Methods:** We used individual-level COVID-19 incidence and mortality data from the U.S. state of Michigan to estimate age-specific incidence and mortality rates by race/ethnic group. Data were analyzed using hierarchical Bayesian regression models, and model results were validated using posterior predictive checks.

**Findings:** In crude and age-standardized analyses we found rates of incidence and mortality more than twice as high than Whites for all groups other than Native Americans. Of these, Blacks experienced the greatest burden of confirmed and probable COVID-19 infection (Age-standardized incidence = 1,644/100,000 population) and mortality (age-standardized mortality rate 251/100,000). These rates reflect large disparities, as Blacks experienced age-standardized incidence and mortality rates 5.6 (95% CI = 5.5, 5.7) and 6.9 (6.5, 7.3) times higher than Whites, respectively. We also found that the bulk of the disparity in mortality between Blacks and Whites is driven by dramatically higher rates of COVID-19 infection across all age groups, particularly among older adults, rather than age-specific variation in case-fatality rates.

**Interpretation:** This work suggests that well-documented racial disparities in COVID-19 mortality in hard-hit settings, such as the U.S. state of Michigan, are driven primarily by variation in household, community and workplace exposure rather than case-fatality rates.

**Funding:** This work was supported by a COVID-PODS grant from the Michigan Institute for Data Science (MIDAS) at the University of Michigan. The funding source had no role in the preparation of this manuscript.

## Introduction

As of September 1, 2020, there have been more than 180K deaths and 6M confirmed and probable cases attributable to SARS-CoV-2 in the United States, with these numbers undoubtedly reflecting a substantial underestimate of the true toll. Geographic, racial-ethnic, age and socioeconomic disparities in mortality have been key features of the first and second wave of the U.S. COVID-19 epidemic.^1–5^ However, the extent to which this differential mortality is driven by evident disparities in rates of infection by age, race, and socioeconomic status, or some combination thereof, remains unknown. Addressing the clear inequities in the toll of death resulting from the COVID-19 pandemic in the U.S. requires disaggregating the relative role of exposure leading to infection versus age-specific case-fatality rates in drivers of the gaping inequity characteristic of SARS-CoV-2 mortality in the United States.

Earlier analyses of other respiratory viruses, such as RSV and influenza, have documented race-ethnic disparities in both rates of infection and case-fatality.^6^ This inequality is driven by diverse factors including comorbid conditions that increase susceptibility to infection as well as disease severity. But it is also a function of structural factors that impact the ability of different groups to avoid infection. Relevant factors include mass incarceration,^7,8^ residential segregation^9,10^ and wealth inequality that facilitates social distancing among the well-off while poorer individuals are more likely to be compelled into ‘essential work’.^11^ A recent cross-national systematic review placed the average infection fatality ratio (IFR) of COVID-19 infection at 0.75%.^12^ However, population age structure is key to shaping the crude mortality rate and total number of deaths from the pandemic, and age-specific variation in the prevalence of underlying conditions is likely to have a dramatic impact on age-specific patterns of mortality.^13^ While some studies have illustrated the differential impact of SARS-CoV-2 on non-White populations in the U.S. using aggregated data,^5^ there are no analyses that provide a clear breakdown of these risks by age, sex, and race.^14^ In the current analysis, we aim to partially close this gap by analyzing patterns of age, sex, and race-specific SARS-CoV-2 infection and mortality using detailed case-level data from the U.S. state of Michigan, which was particularly hard-hit in by SARS-CoV-2 in the winter and spring of 2020, and where the epidemic has been marked by unmistakable racial and socioeconomic iequality.

## Data

We used data from 73,441 people with confirmed and probable COVID-19 infections from the Michigan Disease Surveillance System (MDSS) from March 8^th^, 2020 through July 5^th^, 2020. Each case in MDSS is tagged with a unique identifier, sex at birth, age, race, the date their case was referred to MDHHS, whether the individual died, and the date of death. In order to mitigate the impact of right censored deaths on our case-fatality rate estimates,^15^ we truncated the data at the 97.5% quantile of time to death from case referral date, or forty-six days. This results in truncating all data with a case referral date later than May 20^th^, 2020, after which our data comprise 58,428 individuals.

From this dataset, we excluded 25 cases that did not reside in Michigan or were missing a state of residence, 8,613 people for whom race or ethnicity was not recorded, and 27 people who did not have age recorded or had age > 116 years old indicating entry errors. We combined 68 pairs of records that had duplicate patient identification numbers, resulting in 34 fewer cases. Finally, we dropped 28 patients whose sex at birth was unknown, leading to a final dataset of 49,701 people with a confirmed or probable COVID-19 infection, with known age, race or ethnicity, state of residence, sex at birth, and state prisoner status.

After filtering the individual-level MDSS case data, we binned age by 10 year intervals to age 80, while we combined ages 80 and above in one bin. We also created the race/ethnicity categories of Black/African American, Latino, Asian/Pacific Islander, Native American,Other, and White, where Other comprised the census category of ‘other’, and mixed race people. In order to model per-capita rates of disease we used tract-level population data from the 2010 U.S. Census aggregated to the state level for Michigan to define the population in each age-sex-race stratum.

## Methods

### COVID-19 cumulative indidence rates

To calculate age-specific rates of COVID-19 infection in each age (*i*), sex (*j*), race (*k*), bin, per 100,000 population, we fit a poisson regression model with a population offset term, *log*(*n_ijk_*). where *n_ijk_* is the size of the population for the *ijk*-th group from the 2010 U.S. Census. We included age x sex, age x race, and sex x race interaction terms to capture the full spectrum of potential heterogeneity in our outcome data. We denote the observed number of cases in each group as and the per-capita cumulative incidence rate in each bin as λ_ijk_.

### Case-fatality rates

Age-specific case fatality rates (CFR) were estimated by fitting a binomial model to the number of deaths (*z_ijk_*) as a proportion of the number of total cases (*y_ijk_*) in each age/sex/race bin. We denote the CFR for each group as *ρ_ijk_*, so, *z_ijk_* ∼ Binomial (*y_ijk_*, *ρ_ijk_*). For all analyses of per-capita age-specific incidence rates, we used a log-Gaussian prior distribution with a mean of 0 and standard deviation of 0.1.

### Age and sex standardization

To characterize racial disparities unrelated to differences in population age structure or sex ratios, we employed a post-stratification approach to direct standardization. Specifically, we generated weights for each age x sex bin in the data, using the marginal age and sex distribution for the state of Michigan. We then weighted posterior draws from the incidence and case-fatality models described above to generate adjusted estimates. Age-adjusted case-fatality ratios (CFRs) in our results characterize the ratio in the overall risk of death if individuals in the population were infected at a rate proportional to the share of the total population in their same age/sex bin statewide.

### Software

All analyses were completed in *R* 3.6.3, using the *rstanarm* package^16^ for Bayesian regression analysis, the *tidybayes* package for post-processing^17^ and *ggplot2* for visualization.^18^

## Results

In our dataset, there are 49,701 probable and confirmed COVID-19 cases, and 5,815 deaths attributable to COVID-19, for an overall case-fatality rate of 12%. Of these, 19,662 were among individuals identified as Black or African-American, 23,301 were among individuals identified as White, 1,346 among individuals identified as Asian or Pacific Islander, 123 among individuals identified as Native American, and 1,612 among individuals identified as belonging to any other racial/ethnic group in the 2010 U.S. Census.

Table 1 shows per-capita case and mortality rates by race/ethnic group, as well as corresponding case-fatality rates. Notably, the raw incidence rate among all non-White groups is substantially higher than among Whites for all groups identified in the data except for Native Americans. However, the overall case-fatality rate for Whites is on par with the case-fatality rate for blacks, potentially owing to different distributions of ages among cases and deaths between these groups. Among Whites, the average age of all reported cases was 53.4 yrs (53.2, 53.7), slightly older than among blacks, 51.4 yrs (51.1, 51.6), and significantly older than among Latinos, 38.1 yrs (37.6, 38.6) and those in the “other” race/ethnicity group. For all groups, the mean age among individuals with COVID-19 listed as their cause of death was significantly higher than for all cases within the same group. Among Whites, the average age at death was the greatest, at 79.2 yrs (78.6, 79.9), eight years higher than among Blacks at 71.2 yrs (70.5, 71.9), with Latinos having the youngest average age at death at 66.7 yrs (63.6, 69.8). Taken together, these results suggest that for Non-Whites other than Native Americans, the risk of COVID-19 infection was uniformly higher than for Whites. However, because these groups have differing population age and sex distributions, standardization is necessary to ensure that these reflect differences in risk rather than being a function of the distribution of population.

**Table 1:**
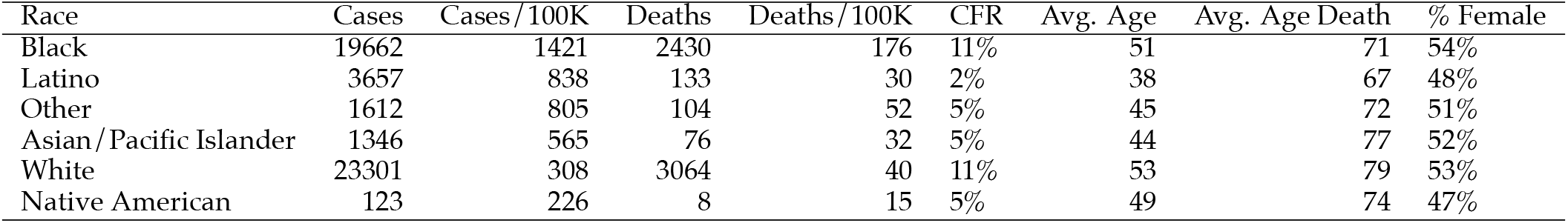
Descriptive statistics of COVID-19 incidence and mortality by race/ethnicity in Michigan, USA, March-June 2020.

| Race | Cases | Cases/100K | Deaths | Deaths/100K | CFR | Avg. Age | Avg. Age Death | % Female |
| --- | --- | --- | --- | --- | --- | --- | --- | --- |
| Black | 19662 | 1421 | 2430 | 176 | 11% | 51 | 71 | 54% |
| Latino | 3657 | 838 | 133 | 30 | 2% | 38 | 67 | 48% |
| Other | 1612 | 805 | 104 | 52 | 5% | 45 | 72 | 51% |
| Asian/Pacific Islander | 1346 | 565 | 76 | 32 | 5% | 44 | 77 | 52% |
| White | 23301 | 308 | 3064 | 40 | 11% | 53 | 79 | 53% |
| Native American | 123 | 226 | 8 | 15 | 5% | 49 | 74 | 47% |

### Standardized incidence and mortality rates

Table 2 shows age and sex-standardized incidence and mortality rates per 100,000 population, and corresponding between-group rate ratios, by race/ethnic group. Rows of the table are ordered by raw incidence per 100K individuals for comparability with Tables 1. This shows that the general patterns in the raw incidence and mortality hold after adjustment, although the age and sex-adjusted incidence among Latinos falls significantly, reflecting the younger average age of cases identified as Latino. The provided incidence rate ratios (IRRs) and mortality rate ratios (MRRs) show the enormous disparity in incidence and mortality between Blacks and Whites, with and IRR of nearly 6 and an MRR of nearly 7. Again, these IRRs and MRRs reflect the fact that all groups other than Native Americans had higher rates of incidence and mortality than whites and that these differences do not simply reflect the age and sex distribution of cases. For Native Americans, rates were statistically indistinguishable from those for Whites, although this may be due to the very small number of cases and deaths overall in this group in our data.

**Table 2:** Age and sex-standardized incidence and mortality rates and corresponding rate ratios. The table shows incidence rates and mortality rates and 95 percent posterior credible inervals, as well as corresponding standardized incidence rate ratios (IRRs) and mortality rate ratios (MRRs). For all comparisons, the incidence and mortality rate among Whites is used as the reference group.

| Race | Incidence/100K | IRR | Mortality/100K | MRR |
| --- | --- | --- | --- | --- |
| Black | 1644 (1621,1668) | 5.6 (5.5,5.7) | 251 (242,262) | 6.9 (6.5,7.3) |
| Latino | 1113 (1074,1152) | 3.8 (3.7,3.9) | 79 (66,94) | 2.2 (1.8,2.6) |
| Other | 1520 (1442,1605) | 5.2 (4.9,5.5) | 152 (124,185) | 4.2 (3.4,5.1) |
| Asian/Pacific Islander | 695 (654,738) | 2.4 (2.2,2.5) | 79 (61,99) | 2.2 (1.7,2.7) |
| White | 293 (289,296) | Ref | 36 (35,38) | Ref |
| Native American | 254 (209,303) | 0.9 (0.7,1) | 26 (12,49) | 0.7 (0.3,1.3) |

### Cumulative incidence rates

Figure 1 shows dramatically higher overall and age-specific incidence rates among Blacks and individuals in the “Other” race/ethnic category than for whites, particularly at older ages at which individuals are far more likely to die from their infection. In addition, the horizontal dashed line in each panel of Figure 1 shows the raw incidence rate for each group, and then dotted line indicates the age-adjusted incidence rate.

**Figure 1:**
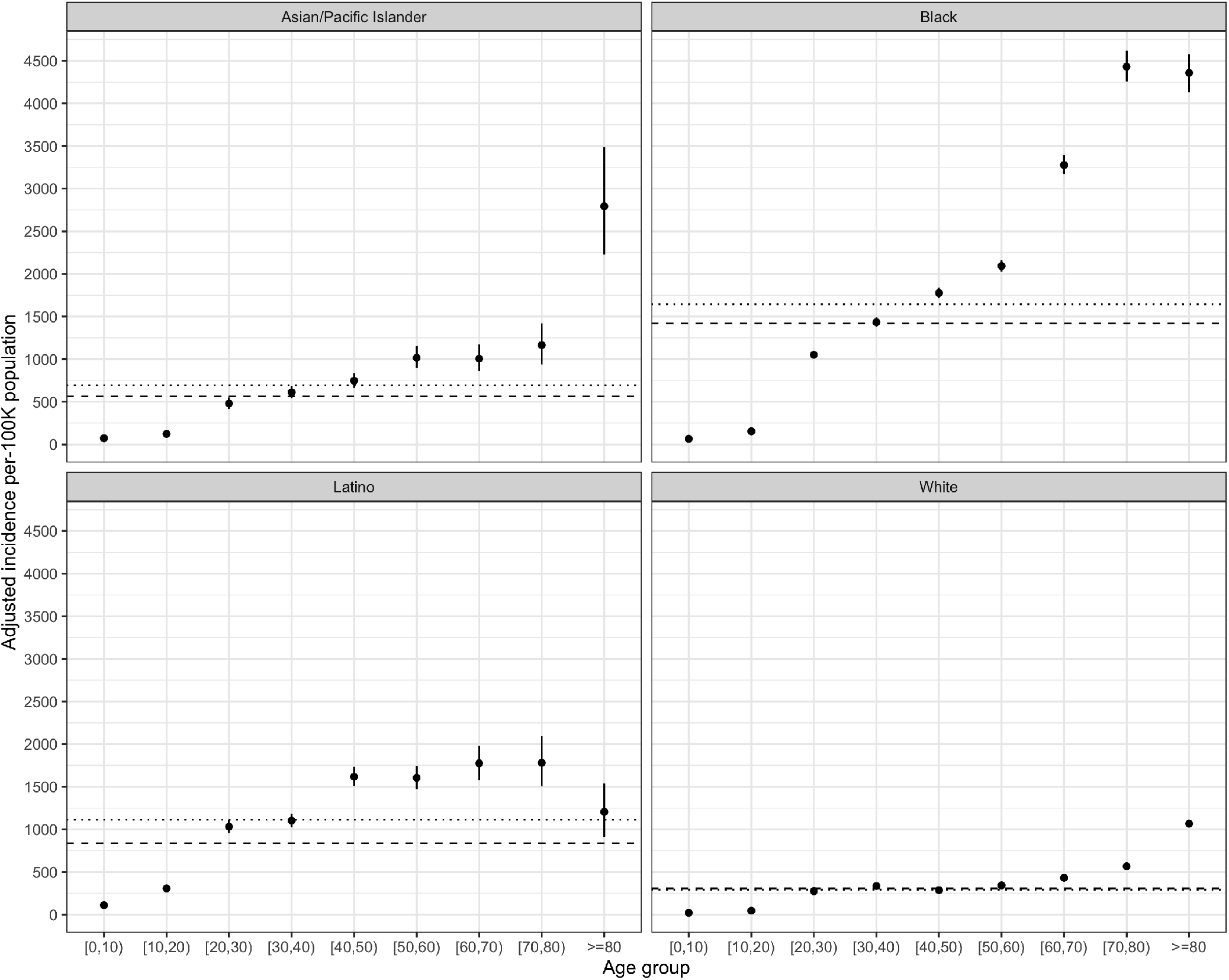
Incidence of COVID-19 infection per 100K population by age and race. Dashed lines indicate the crude overall rate for each group, dotted lines indicate group-specific age and sex-adjusted rates.

The extent of these disparities in incidence is clearly in evidence in panel A of Figure 3, which shows the ratio of the age-specific cumulative incidence rate (incidence rate ratio, IRR) for each race/ethnic group as compared to the comparable rate for Whites. In this case, rates for all non-White groups are significantly higher, with these disparities most pronounced at older ages for Blacks, and younger ages for latinos. The IRR for individuals in the ‘other’ group was fairly consistent across ages, with a small drop in the 20-40 age range.

In the following sections, we will examine age-stratified incidence and mortality rates by race/ethnicity for Blacks, Latinos, Asians/Pacific Islanders and Whites. Native Americans are excluded from age-stratified analyses due to a small sample size, as are individuals in the “Other” race/ethic categorization.

### Case-fatality rates

Figure 2 illustrates the trend in case-fatality rates by age. These are fairly consistent between groups, with a steadily increasing trend in the probability of death among identified cases from age 50 onwards, although there are apparent differences in these rates at younger ages as well. These are clearly visible in panel B of Figure 3, which shows the ratio of the age-specific case-fatality ratio for Blacks, Latinos and those in the ‘other’ group vs. the case-fatality rate for Whites of the same age. Because of the small number of deaths in individuals < 20 years of age, these groups are excluded from the figure. For Blacks, all age groups from 30 to 70 years experience higher case-fatality rates than Whites, with this disparity most pronounced in the 40-49 year age-group. However, for Latinos and those in the ‘other’ race/ethnic group, there are no significant differences in age-specific case-fatality rates as compared to Whites. In Figs. 2 & 3, the dashed line indicates raw case-fatality rates, while the dotted line indicates an age-adjusted estimate of case-fatality assuming an equal probability of infection at all ages of the standardized population. These results suggest that although there are meaningful differences in case-fatality by race and age, that the large raw and standardized disparities in COVID-19 mortality cannot be explained by case fatality rates alone.

**Figure 2:**
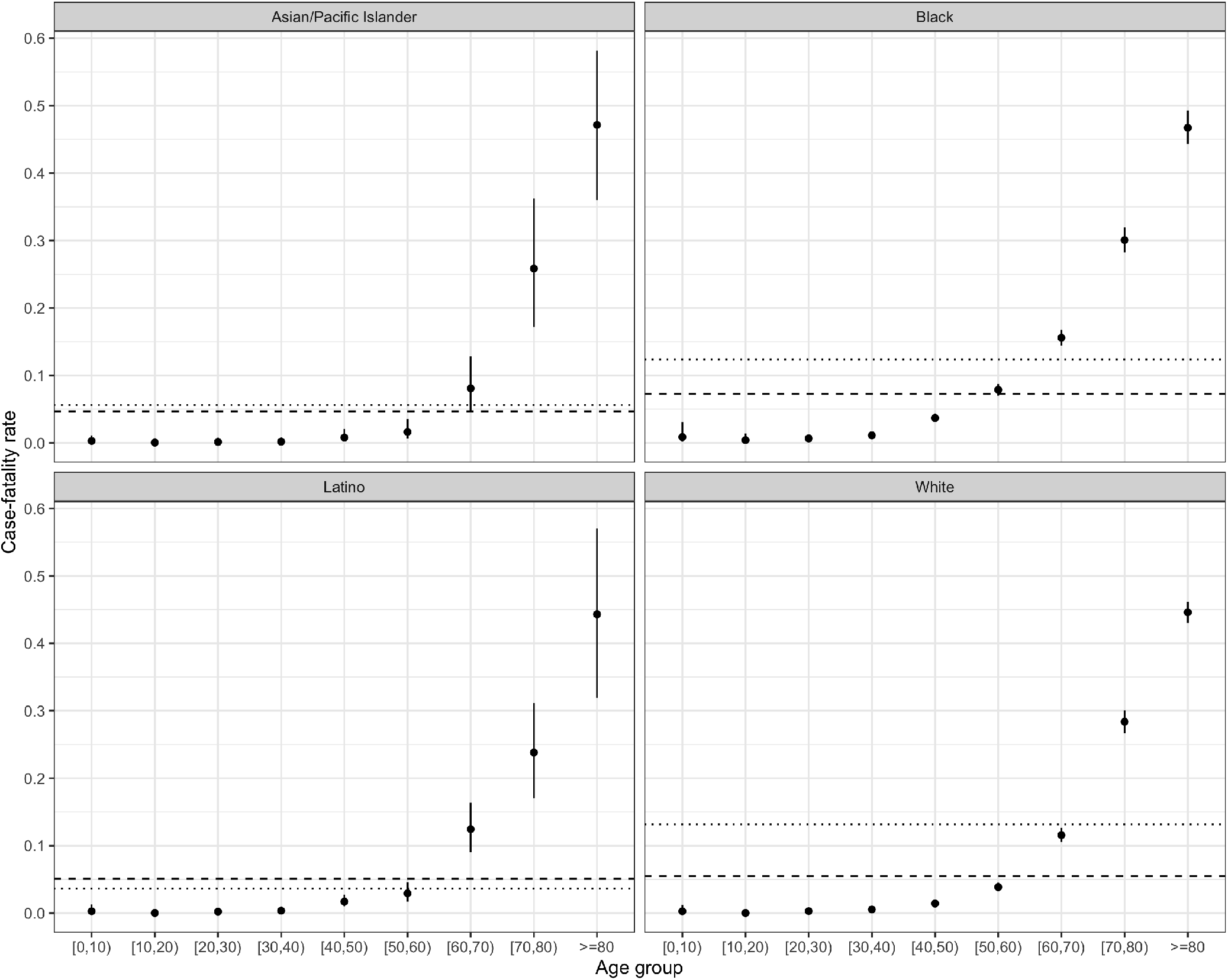
COVID-19 case-fatality rate by age and race. Dashed lines indicate the crude overall rate for each group, dotted lines indicate group-specific age and sex-adjusted rates.

**Figure 3:**
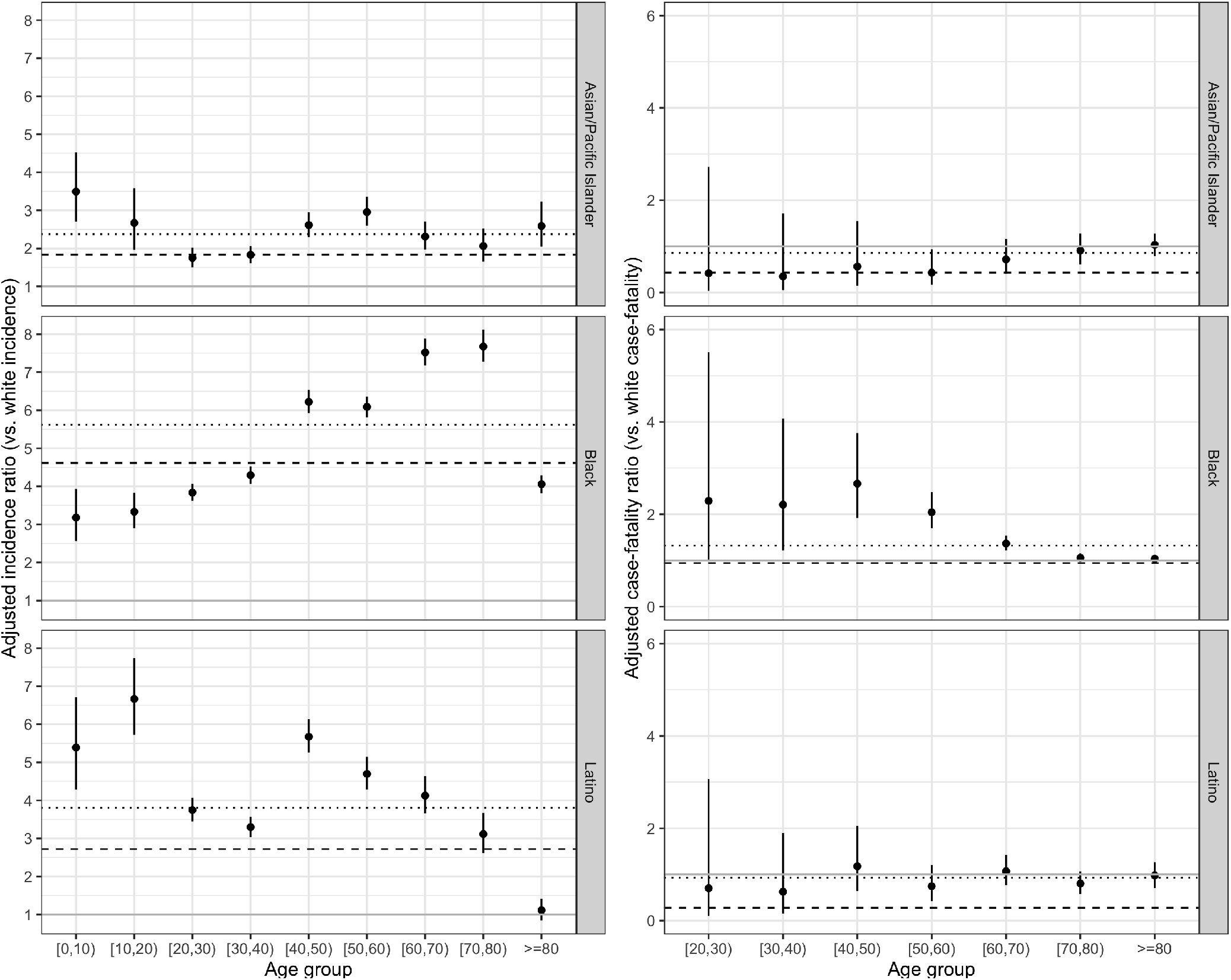
Disparities in COVID-19 incidence and case-fatality rate by age and race. Dashed lines indicate the ratio in the crude overall rate for each group, dotted lines indicate group-specific age and sex-adjusted rate ratios. The solid gray line is a guide for assessing the strength of association.

One way to understand the relative importance of exposure vs. case-fatality on the disparate burden of mortality by race is to examine the counterfactual scenario in which each race/ethnic group has the age- and sex-specific COVID-19 incidence rate of the corresponding age/sex group among Whites but their original age- and sex-specific case-fatality rates. When we do this, we find that this would result in a decrease of 83%, 95% CI = (82%,84%) of deaths among Blacks, 63%, 95% CI = (53%,72%) among Latinos, and 59%, 95% CI = (46%,69%) among Asian/Pacific Islanders.

These results suggest that while differential case-fatality rates can account for some of the disparity in Black vs. White mortality rates, the large majority of COVID-19 deaths among African-Americans in Michigan can be attributed to the large differences in age-specific incidence illustrated in Fig 1. Similarly, although Latinos and Asian/Pacific Islanders have similar crude mortality rates to Whites (Table 1), these results indicate that these rates would be significantly lower if their exposure risks were more similar to their White peers.

Another potential explanation for racial differences in crude incidence and mortality is that there may be differential risks by sex that vary over race/ethnic categories. However, the age-standardized results in Table 2 suggest this is unlikely to be the case. We also explicitly examined sex-specific differences in incidence, overall mortality and case-fatality (See Figure S1 in the supplementary materials), and found no meaningful differences by sex or race, although the risk of death from COVID-19 is significantly higher for men than for women across all groups, echoing other findings.

## Discussion

Our results highlight yawning gaps in COVID-19 incidence and mortality in Michigan that cannot be explained away by differences in population age and sex composition. Our results also suggest that the stark differences in crude and adjusted mortality between Blacks and all other race/ethnic groups shown in Tables 1 & 2 are driven in large part by disparities in infection risk at all ages, and an extremely high rate of COVID-19 infection among older Blacks in particular. This group of individuals had a similar case-fatality rate to same-aged Whites, but reported infection rates 6-8 times greater than among their White counterparts. Some of this disparity is also driven by the higher case-fatality rate among middle-aged Blacks, as compared to same-aged Whites, in combination with the 5-6 times greater risk of infection among middle-aged African-Americans as compared to Whites.

Despite these unambiguous results, the full extent of racial and socioeconomic disparities in COVID-19 outcomes in Michigan and the rest of the U.S. is likely to be even wider than is reflected in administrative data of the type analyzed here. Results from other hard-hit cities, states, and countries have indicated high rates of excess mortality reflective of unrecognized and unreported COVID-19 infection.^19^ In a recent analysis using state-level data Weinberger et al.^20^ estimated rates of excess mortality due to COVID-19. They found that there were approximately 4700 unreported deaths likely due to COVID-19 or another respiratory infection during the period from March 1 – May 30 2020, for a rate of 61/100,000 unreported deaths from COVID-19 above the reported totals. Of course, the damage to health from the pandemic is not only among infections and deaths from SARS-CoV-2: In other recent results, Woolf et al.^21^ showed that 33% of the total excess deaths during the period from March 1-April 25 2020 in Michigan were attributable to non-infectious causes, with the remainder associated with respiratory infections, primarily COVID-19. Though these results are not broken down by race/ethnicity, it remains critically important to understand who is among these unreported deaths from COVID-19 as well as non-infectious illnesses. Beyond the delay in healthcare seeking of all risk averse individuals, it is quite likely that these patterns of excess death reflect underlying disparities in chronic illnesses that predispose individuals to mortality from COVID-19, lack of access to healthcare for Blacks, Latinx individuals and other minority groups, and variable quality of care delivered based on racial-ethnic identity.

When interpreting these and other results illustrating racial disparities in COVID-19 incidence and outcomes, it is key to not examine race as a risk factor independent of health conditions, wealth and other potentially modifiable risk factors^22^ that may predispose individuals to COVID-19 infection and mortality. For example, McClure et al.^23^ illustrate how a focus on – and adjustment for – individual-level “underlying conditions” obscures the role of racial inequality in shaping the prevalence of these chronic health conditions, and other factors such as intergenerational households, which may increase risk among racial and ethnic minority groups.

A strength of our analysis is the use of detailed case data obtained directly from the Michigan Disease Surveillance System. This allowed us to identify age and race-specific risks of COVID-19 infection and death. Nonetheless, there are some limitations that are important to highlight. First, our reliance on census-defined race/ethnicity as a proxy for exposure and mortality risk is necessarily reductive and does not shed enough light factors that can be modified to reduce these disparities. Numerous studies have highlighted the role of wealth and other markers of socioeconomic status (SES) such as educational attainment, as an important mediator of the effects of race on health outcomes. At the same time, even after adjustment for SES, there is often a residual effect of race which cannot be explained solely in terms of material wealth, and instead is likely accounted for by other factors including discrimination in healthcare settings, and the impact of cumulative stress associated with exposure to structural racism.^24^ Future analyses are necessary using either prospectively collected data inclusive of SES, or spatial analyses that join neighborhood-level information on wealth and other markers of SES with individual-level case data.

In addition, although the disparities in our data likely mirror many of those nationwide, it is important to remember that these results reflect specifically patterns of infection and death in Michigan during the first wave of the COVID-19 pandemic in the U.S. Although its relatively large population size and socioeconomic and racial composition make the state a belwether of many national trends, patterns of racial residential segregation are more regional than in other hard-hit locales, such as New York City, Chicago, Los Angeles, and Atlanta.^25^ While there is a substantial Latino and Asian population in Michigan, patterns of racial and socioeconomic diversity are not directly comparable to those of key centers of ongoing transmission such as Florida, Texas, and California. Consequently, similar analyses from these contexts are urgenly needed.

Another important limitation of our results is the reliance on racial categorizations from the 2010 census as well as the documentation of race/ethnicity on case reports and death certificates. For example, the one exception to the pattern of disparate risks among non-whites was the finding of no difference in incidence and mortality risk for Native Americans in Michigan as compared to whites. These results may reflect the concerted effort to prevent transmission in Native communities.^26^ However, this may also reflect the fact that cases among American Indians and Alaska Natives have disproportionately been categorized as occurring among the “Other” race/ethnic groups.^27^ Other important groups are not included in census statistics. For example, there is a large Arab-American population in Southeast Michigan,^28^ who experience many social and medical vulnerabilities that may put them at increase risk for COVID-19 infection and death.^29^ However, the lack of available census data and uniform reporting of Arab ethnicity in medical records precludes rigorous analysis of COVID-19 infection and mortality risks among this important group.

Because of the deep structural roots of the disparities identified in this analysis, it is easy – but wrongheaded – to conclude that there is nothing to be done. The fluid nature of the COVID-19 pandemic and its response provides opportunities to slow the tide of transmission and death. For this to be the case, however, similar amounts of effort to what is being done to ensure that college campuses and other workplaces can re-open safely needs to be focused on increasing the quality and quantity of testing, healthcare and social support among people of color. Further, there is a need to examine the racialized dismantaling of public infrastructure and systematic divestment which drives disparities in both exposure, susceptibility and mortality.^30^ However, while understanding the causes of disparate outcomes is important, it does not necessarily instruct us on what to do next. If the current pandemic teaches us something, it is that closing the gap in infection and mortality during the current catastrophe – and preventing such inequities in the next one – requires a re-orientation around an ‘epidemiology of consequence’^31^ that is focused on identifying strategies to attack the structural and practical barriers to health equity before the next disaster strikes.

## Data Availability

Because the underlying data are identifiable, they are not available publicly. Summaries of data are available at https://www.covidmapping.org

## Author Contributions

JZ, RT conceptualized and completed data analysis; JZ, RT, RN, AC, NM, PD participated in data preparation and cleaning; all authors participated in the writing and editing of the final manuscript.

## Acknowledgments

The authors gratefully acknowledge the efforts and insight of Sarah Lyon-Callo, Jim Collins, and other members of the Michigan Dept. of Health and Human Services, as well as the input of Emily Martin, Marisa Eisenberg and Sharon Kardia at the University of Michigan.

## Funding

This work was supported by a *COVID-PODS* grant from the Michigan Institute for Data Science (MIDAS) at the University of Michigan and the *MCubed* program at the University of Michigan. The funding sources had no role in the preparation of this manuscript.

## Supplementary Materials

### Supplementary Results

**Figure S1:**
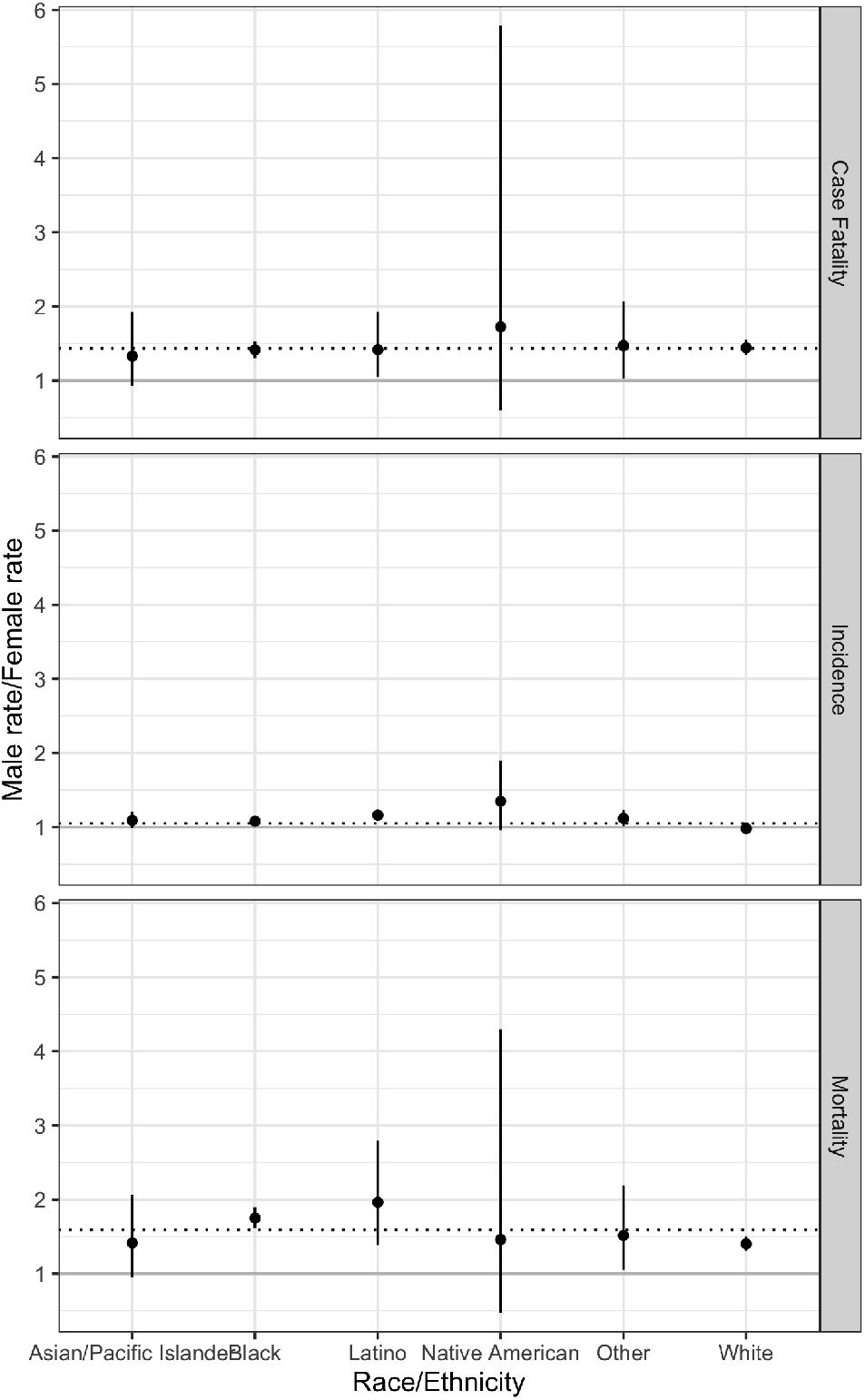
Male vs. female risk ratios for key COVID-19 risks.

### Posterior Predictive Checks

**Figure S2:**
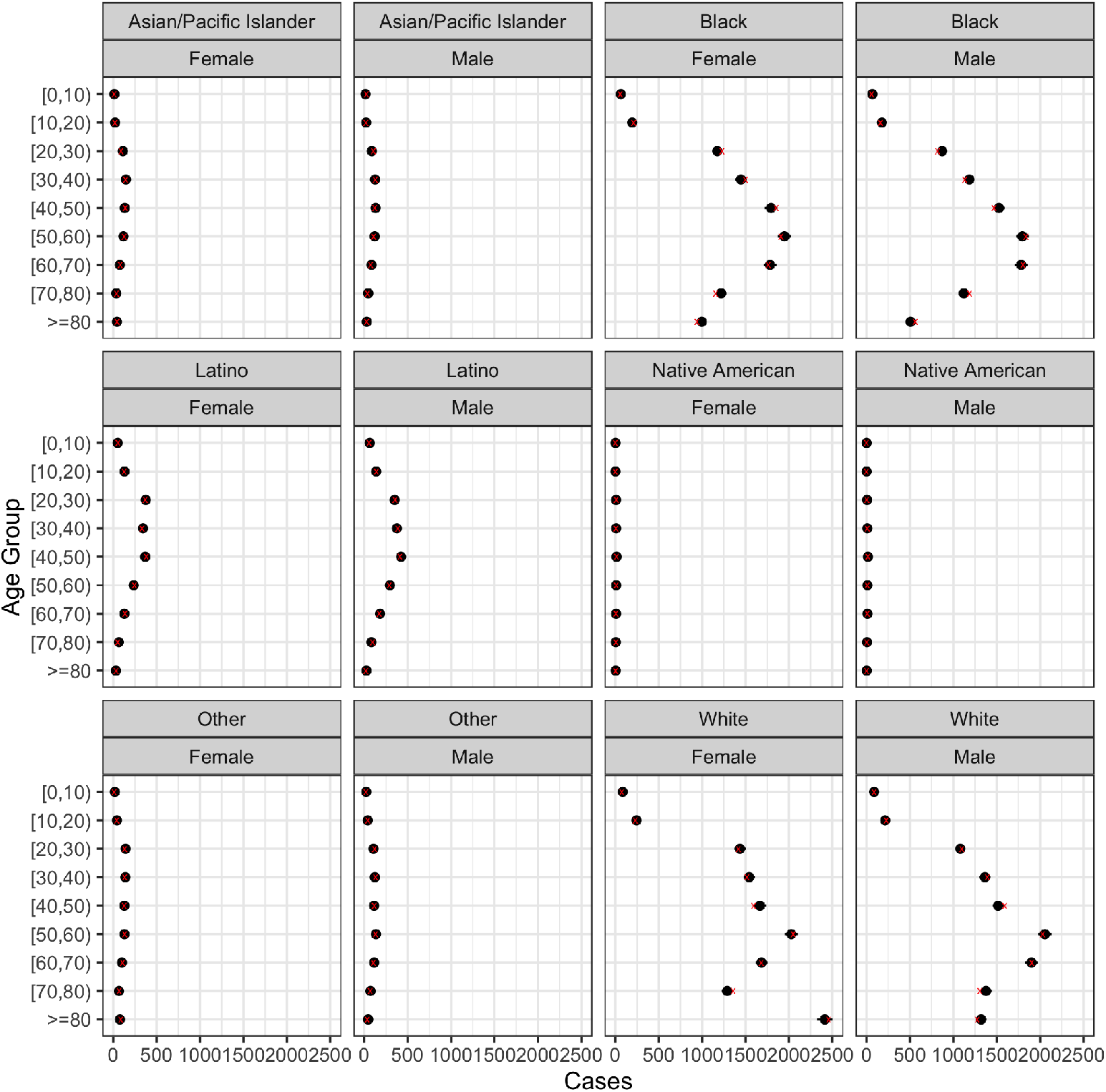
Posterior predictive check for number of incident cases. Circles indicate predicted counts, x indicates raw counts.

**Figure S3:**
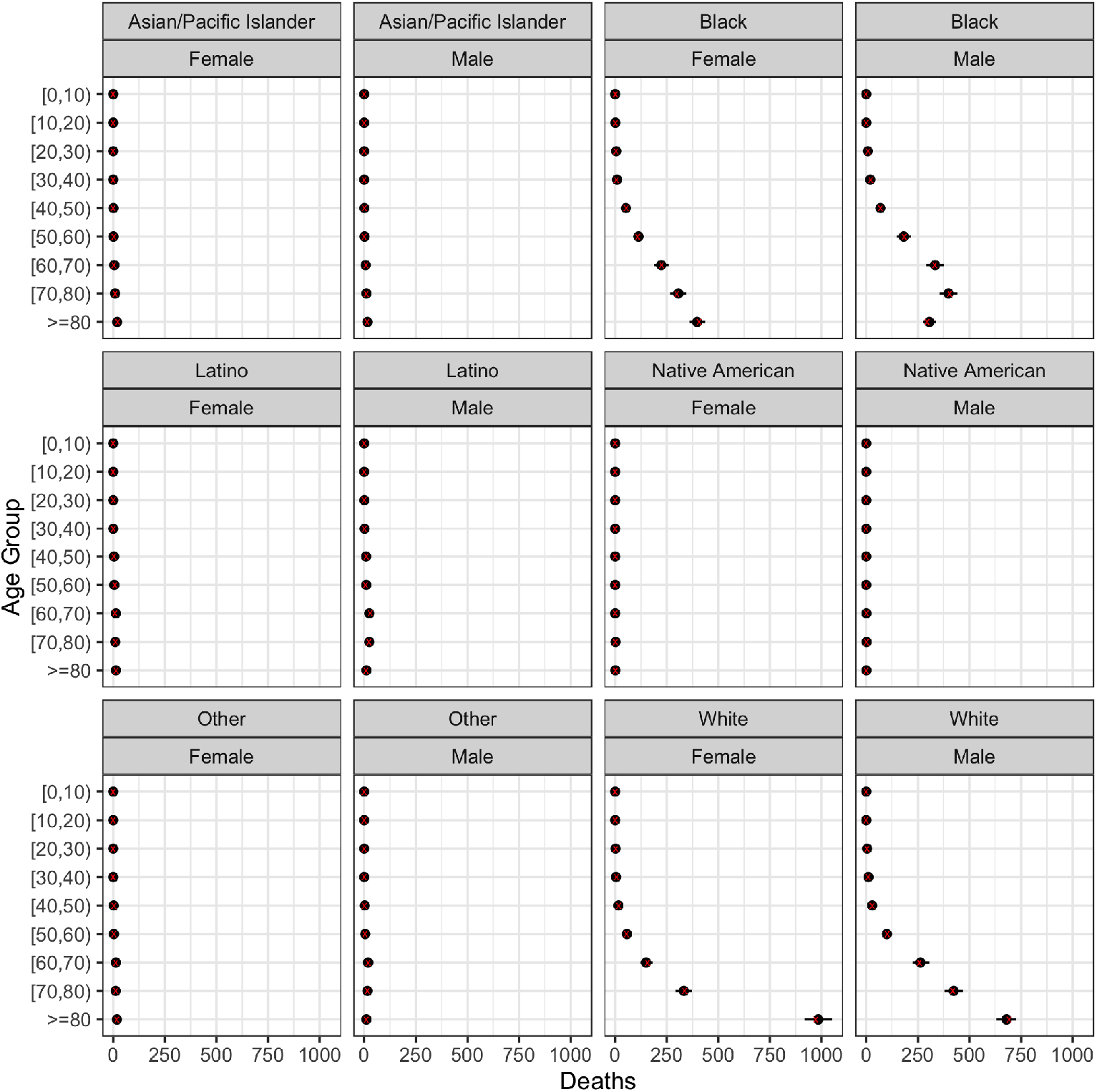
Posterior predictive check for number of deaths. Circles indicate predicted counts, x indicates raw counts.

## Notes

### Competing Interest Statement

The authors have declared no competing interest.

### Author Declarations

All analyses were approved by the University of Michigan Health Sciences IRB and the Michigan Department of Health and Human Services IRB

